# Cardiorespiratory fitness, polygenic risk, and breast cancer in postmenopausal women: a prospective cohort study

**DOI:** 10.64898/2026.03.12.26347589

**Authors:** Kumpei Tanisawa, Daiki Watanabe, Qian Li, Xiayue Fan, Xiaomin Sun

**Affiliations:** Faculty of Sport Sciences, Waseda University, Tokorozawa, Saitama 359-1192, Japan; National Institute of Biomedical Innovation, National Institutes of Biomedical Innovation, Health and Nutrition, Ibaraki-city, Osaka, Japan; Med-X Institute, Center for Immunological and Metabolic Diseases, the First Affiliated Hospital of Xi’an Jiaotong University, Xi’an Jiaotong University, Xi’an, Shaanxi 710049, China; Global Health Institute, School of Public Health, Xi’an Jiaotong University Health Science Center, Xi’an, Shaanxi 710061, China

**Keywords:** Breast cancer, Cardiorespiratory fitness, Polygenic risk score, UK Biobank, Prospective cohort

## Abstract

**Purpose:** Although cardiorespiratory fitness (CRF) is inversely associated with breast cancer risk, whether CRF modifies the association between breast cancer polygenic risk score (PRS) and incident breast cancer remains unclear. We examined the independent and joint associations of CRF and breast cancer PRS with incident breast cancer in postmenopausal women from the UK Biobank and evaluated multiplicative and additive interactions between CRF and breast cancer PRS.

**Methods:** This prospective cohort study included postmenopausal women from the UK Biobank. CRF was estimated using a submaximal cycle ergometer test, and genetic susceptibility was assessed using a breast cancer PRS. Cox proportional hazards models adjusted for established breast cancer risk factors were used to estimate hazard ratios. Multiplicative and additive interactions between CRF and PRS were evaluated, with additive interaction assessed using the relative excess risk due to interaction (RERI).

**Results:** Among 15,199 postmenopausal women, 547 women developed incident breast cancer during a median follow-up of 10.7 years. Higher CRF was associated with lower risk of incident breast cancer in a dose–response manner, whereas higher PRS was associated with higher risk. Although multiplicative interaction was not significant, additive interaction analyses indicated that higher CRF was associated with attenuation of excess breast cancer risk conferred by high PRS (RERI −0.79, 95% CI −1.47 to −0.10). This attenuation was particularly evident among women aged ≥60 years.

**Conclusions:** Higher CRF was associated with lower breast cancer risk and attenuation of excess breast cancer risk conferred by elevated polygenic risk, particularly among older women. These findings support the potential role of CRF in risk-stratified breast cancer prevention strategies.

## Introduction

Breast cancer is the most commonly diagnosed cancer among women worldwide and remains a major public health concern because of its high incidence and disease burden (1). Although advances in screening and treatment have improved survival, primary prevention through modification of risk factors remains an important public health priority (2).

In addition to established lifestyle-related risk factors, genetic susceptibility plays a substantial role in breast cancer development (3). Genome-wide association studies (GWASs) have identified numerous common genetic variants associated with breast cancer risk (4, 5). These discoveries have led to the development of polygenic risk scores (PRSs), which capture the combined effects of these variants and enable breast cancer risk stratification across the general population (6, 7). Individuals with elevated polygenic risk have substantially greater risk of developing breast cancer than those with lower polygenic risk (6), highlighting the importance of identifying modifiable factors that may attenuate this excess risk.

Cardiorespiratory fitness (CRF) is an objective physiological measure reflecting the integrated capacity of the cardiovascular and respiratory systems and is influenced by habitual levels of physical activity as well as genetic and physiological factors (8). Because of its strong and consistent associations with morbidity and mortality (9, 10), CRF has been widely recognized as an important clinical indicator of overall health and disease risk. Prospective cohort studies have demonstrated an inverse dose–response association between CRF and incident breast cancer (11, 12). Considering the established clinical importance of CRF, clarifying whether higher CRF is associated with attenuation of the excess breast cancer risk conferred by elevated polygenic risk may contribute to optimizing breast cancer prevention strategies. However, whether higher CRF modifies the association between polygenic risk, as quantified by PRS, and incident breast cancer remains unclear.

Therefore, the present study aimed to examine the independent and joint associations of CRF and breast cancer PRS with incident breast cancer in postmenopausal women from the UK Biobank. We further evaluated multiplicative and additive interactions between CRF and breast cancer PRS to assess whether CRF modifies the association between polygenic risk and incident breast cancer.

## Methods

### Study Population

The UK Biobank is a large prospective cohort study that recruited over 500,000 participants aged 37–73 years between 2006 and 2010 across the United Kingdom (13). At baseline, participants completed detailed questionnaires, underwent standardized physical examinations, and provided biological samples.

For the present analysis, we included postmenopausal women with available CRF and breast cancer PRS data. Premenopausal women were excluded because of the difference in breast cancer etiology. Postmenopausal status was defined as natural menopause or menopause resulting from bilateral oophorectomy. Participants were excluded if they were men (*n* = 228,973); lacked estimated maximal oxygen uptake (V O max) data (*n* = 238,082); were premenopausal at baseline (*n* = 12,198); had a history of any cancer at baseline (*n* = 2,736); were of non-European genetic ancestry (n = 4,722), based on UK Biobank data-field 22,006 (Genetic ethnic grouping); had missing PRS data (*n* = 65); or had extreme V O max values (≥3 standard deviations from the mean, *n* = 153). The final analytical sample comprised 15,199 postmenopausal women (Figure 1), all of whom were classified as White British based on self-reported ethnic background (UK Biobank data-field 21000).

**Figure 1.**
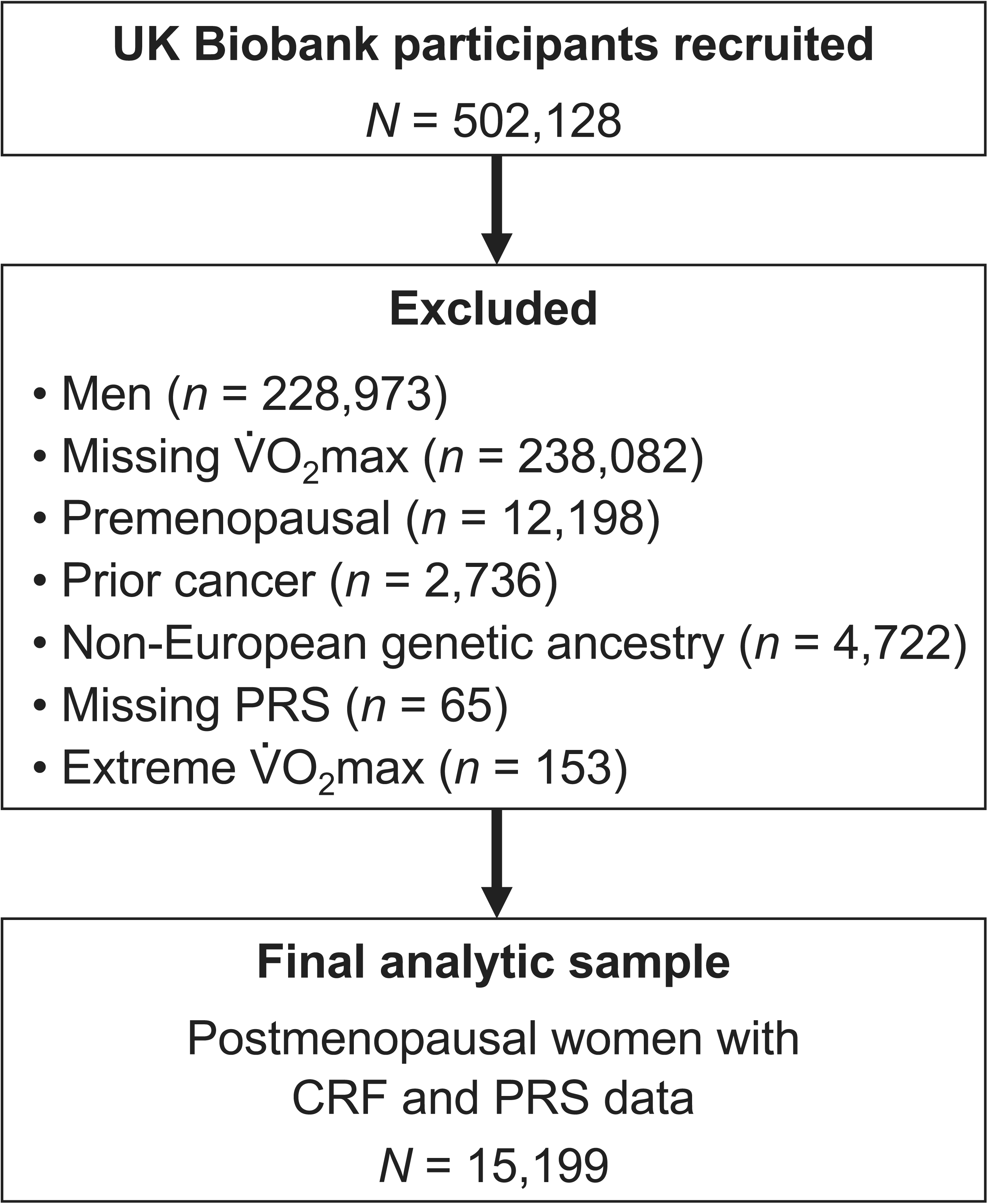
Flow diagram of participant selection from the UK Biobank. The final analytic sample comprised 15,199 postmenopausal women with available CRF and PRS.

### Assessment of CRF

CRF was assessed by estimating V O max (mL·kg ¹·min ¹) using a validated submaximal cycle ergometer test administered at baseline. V O max was derived from heart-rate responses during graded submaximal exercise, incorporating age, sex, body mass, and workload, according to the UK Biobank protocol (14). This procedure has been validated against directly measured V O_2_max (15). To account for age-related differences in CRF, participants were categorized into age-specific quartiles of V O max within predefined age groups (40–49 years, 50–59 years, and ≥60 years) for categorical analyses. Age-specific quartile cut-points for CRF are provided in Supplemental Table 1 (Supplemental Digital Content 1).

### Breast Cancer PRS

Genetic susceptibility to breast cancer was assessed using the breast cancer PRS, previously developed using external GWAS data and available in the UK Biobank (data-field 26,220) (16). The PRS was calculated as the genome-wide sum of per-variant posterior effect sizes weighted by allele dosage, followed by centering and variance standardization by genetic ancestry. Participants were categorized into quartiles of the PRS for categorical analyses. Quartile cut-points for PRS are provided in Supplemental Table 1 (Supplemental Digital Content 1).

### Ascertainment of Breast Cancer

Incident breast cancer cases were ascertained through linkage to national cancer registries of England, Scotland, and Wales and defined as the first primary diagnosis of breast cancer occurring after the date of baseline assessment, identified using International Classification of Diseases, 10th Revision (ICD-10) codes C50. Participants were followed from the date of baseline assessment until breast cancer diagnosis, death, loss to follow-up, and the end of follow-up, whichever occurred first. Because all participants included in the present analysis attended assessment centers in England, follow-up was censored on 31 December 2020, corresponding to the cancer registry follow-up end date for England. As a secondary outcome, breast cancer–specific mortality was defined as death with breast cancer (ICD-10 code C50) recorded as the underlying cause, using the same ascertainment procedures described above.

### Covariates

Covariates were selected a priori based on established breast cancer risk factors. These included family history of breast cancer (yes, no), socioeconomic status assessed using the Townsend deprivation index (quintile), alcohol consumption frequency (<3 times a month, 1–4 times a week, daily or almost daily), smoking status (never, former, current), ever use of hormone replacement therapy (yes, no), age at menarche (<12, 12–13, ≥14 years), number of live births (0, 1, 2, ≥3 live births), age at first birth (<25, 25–29, ≥30 years), and age at menopause (<45, 45–49, 50–54, ≥55 years, Unknown). Age at menopause was based on the reported age at natural menopause. When menopause was attributable to oophorectomy and the age at oophorectomy was available, the age at oophorectomy was used. Women for whom age at menopause was structurally unavailable were classified as Unknown. Family history of breast cancer was defined using self-reported maternal and sibling history. Family history was included because it may capture familial risk not fully explained by the PRS. Body fat percentage (continuous) was considered an adiposity-related covariate. All covariates were defined using measurements and questionnaire responses collected at baseline. BMI, which was used for subgroup analyses, was calculated as weight in kilograms divided by height in meters squared (kg/m²), using height and weight measured during the baseline assessment at the UK Biobank assessment center.

### Statistical Analysis

All analyses were conducted using R version 4.5.1, with statistical significance set at *P* < 0.05. Missing covariate data were handled using multiple imputation by chained equations (20 datasets) under the missing-at-random assumption. The proportion of missing data for each covariate is shown in Supplemental Table 2 (Supplemental Digital Content 1). Imputation was applied only to covariates with missing data; primary exposures (CRF and PRS) and the outcome were not imputed. The imputation model included the exposures, covariates, and the breast cancer event indicator. For age at menopause, ordinarily missing values were multiply imputed, whereas structurally unavailable values were not imputed and were retained as Unknown. Models were fitted separately within each imputed dataset. Regression coefficients and their variances were pooled using Rubin’s rules, whereas likelihood ratio test statistics for multiplicative interaction were combined across imputations using the D2 method. Associations of CRF and PRS with incident breast cancer and breast cancer–specific mortality were examined using Cox proportional hazards models with attained age as the underlying time scale. The proportional hazards assumption was evaluated using Schoenfeld residuals, and no meaningful violations were observed. Model 1 used attained age as the underlying time scale and included no covariates beyond the exposure of interest. Model 2 was adjusted for established breast cancer risk factors (family history of breast cancer, Townsend deprivation index, alcohol consumption frequency, smoking status, hormone replacement therapy use, age at menarche, number of live births, age at first birth, and age at menopause). Because adiposity may act both as a confounder and as a potential mediator of the association between CRF and breast cancer risk, Model 3 was additionally adjusted for body fat percentage to examine whether the association between CRF and breast cancer risk was independent of adiposity. When PRS was the primary exposure, Models 1 and 2 were applied. Hazard ratios (HRs) and 95% confidence intervals (CIs) were estimated across quartiles of CRF and PRS using the lowest quartile as the reference. CRF and PRS were also analyzed as continuous variables (per 1 metabolic equivalent of task (MET), equivalent to 3.5 mL·kg□¹·min□¹, and per SD increase, respectively), and statistical significance was assessed using Wald tests. Nelson–Aalen cumulative hazard curves were constructed to visualize the cumulative hazard of breast cancer across CRF and PRS categories. Restricted cubic spline regression was used to examine potential non-linear associations of CRF and PRS with breast cancer risk. For the joint association analysis of CRF and PRS, participants were dichotomized into low-to-moderate (Q1–Q3) and high (Q4) categories for both CRF and PRS. Cox proportional hazards models were then fitted using the joint category of low-to-moderate PRS and low-to-moderate CRF as the reference group. Multiplicative interaction between CRF and PRS categories was evaluated by including cross-product terms, with statistical significance assessed using likelihood-ratio tests. Additive interaction between CRF and PRS categories was evaluated using the relative excess risk due to interaction (RERI), with 95% CIs derived using the delta method and pooled across imputations. Subgroup analyses were performed according to age (40–59 and ≥60 years) and BMI (<25 and ≥25 kg/m²). BMI was selected for subgroup analyses because BMI-defined overweight and obesity categories are widely used in clinical practice and breast cancer epidemiology. Sensitivity analyses excluded breast cancer cases diagnosed within the first 2 years of follow-up to reduce the potential for reverse causation. As a sensitivity analysis accounting for competing risks, Fine–Gray subdistribution hazard models were fitted for incident breast cancer, treating all-cause death before breast cancer diagnosis as a competing event.

## Results

### Participant Characteristics

During a median follow-up of 10.7 years (IQR 10.6–10.8), 547 incident breast cancer cases were identified among 15,199 postmenopausal women. Table 1 presents the baseline characteristics of the study population according to quartiles of CRF before multiple imputation. Women in higher CRF quartiles had lower body fat percentage and more favorable lifestyle profiles compared to those in lower CRF quartiles.

**Table 1.** Characteristics of participants at baseline according to CRF quartiles.

|  | Total | CRF quartiles |  |  |  |
| --- | --- | --- | --- | --- | --- |
|  |  | Q1 | Q2 | Q3 | Q4 |
| *n* | 15199 | 3835 | 3859 | 3776 | 3729 |
| Age at baseline (years) | 61.0 (57.0, 64.0) | 61.0 (57.0, 65.0) | 61.0 (57.0, 65.0) | 61.0 (57.0, 64.0) | 61.0 (56.0, 64.0) |
| Height (cm) | 163.0 (159.0, 167.0) | 162.0 (158.0, 166.0) | 163.0 (159.0, 167.0) | 163.0 (159.0, 167.0) | 163.0 (159.0, 167.0) |
| Body mass (kg) | 68.8 (61.7, 77.5) | 79.4 (70.7, 89.4) | 71.3 (65.0, 78.7) | 66.8 (61.3, 72.7) | 61.4 (56.5, 66.7) |
| BMI (kg/m<sup>2</sup>) | 25.9 (23.4, 29.2) | 30.1 (26.9, 33.8) | 27.0 (24.8, 29.7) | 25.2 (23.3, 27.4) | 23.2 (21.6, 25.0) |
| Body fat percentage (%) | 37.0 (32.4, 41.3) | 42.2 (38.2, 45.8) | 38.5 (34.8, 41.9) | 35.7 (32.2, 39.0) | 31.9 (28.2, 35.4) |
| Estimated V<sub>O2</sub>max (mL·kg<sup>-1</sup>·min<sup>-1</sup>) | 24.4 (21.5, 27.7) | 19.4 (17.6, 20.6) | 23.0 (22.2, 23.8) | 25.9 (24.9, 26.8) | 30.3 (28.7, 32.5) |
| Breast cancer PRS | -0.18 (-0.84, 0.51) | -0.19 (-0.84, 0.51) | -0.19 (-0.84, 0.51) | -0.16 (-0.84, 0.55) | -0.19 (-0.85, 0.47) |
| Family history of breast cancer | 1718 (11.4) | 460 (12.1) | 409 (10.7) | 427 (11.4) | 422 (11.4) |
| Townsend deprivation index | -2.24 (-3.63, 0.01) | -2.10 (-3.55, 0.24) | -2.21 (-3.60, -0.13) | -2.31 (-3.65, -0.12) | -2.30 (-3.71, 0.04) |
| Alcohol consumption frequency |  |  |  |  |  |
| <3 times a month | 5089 (33.5) | 1659 (43.3) | 1354 (35.1) | 1122 (29.8) | 954 (25.6) |
| 1, 4 times a week | 7188 (47.3) | 1666 (43.5) | 1812 (47.0) | 1860 (49.3) | 1850 (49.6) |
| Daily or almost daily | 2913 (19.2) | 509 (13.3) | 691 (17.9) | 789 (20.9) | 924 (24.8) |
| Smoking status |  |  |  |  |  |
| Never | 8980 (59.3) | 2309 (60.5) | 2356 (61.1) | 2168 (57.6) | 2147 (57.7) |
| Former | 5165 (34.1) | 1267 (33.2) | 1252 (32.5) | 1334 (35.5) | 1312 (35.2) |
| Current | 1009 (6.7) | 241 (6.3) | 246 (6.4) | 259 (6.9) | 263 (7.1) |
| Ever use of hormone replacement therapy | 7349 (48.4) | 1813 (47.4) | 1923 (49.9) | 1899 (50.4) | 1714 (46.1) |
| Age at menarche |  |  |  |  |  |
| <12 | 3062 (20.7) | 958 (25.7) | 790 (21.0) | 680 (18.5) | 634 (17.6) |
| 12–13 | 6566 (44.4) | 1603 (43.0) | 1672 (44.4) | 1669 (45.4) | 1622 (45.0) |
| ≥14 | 5154 (34.9) | 1167 (31.3) | 1307 (34.7) | 1328 (36.1) | 1352 (37.5) |
| Number of live births |  |  |  |  |  |
| 0 | 2782 (18.3) | 694 (18.1) | 671 (17.4) | 681 (18.0) | 736 (19.8) |
| 1 | 1852 (12.2) | 443 (11.6) | 495 (12.8) | 500 (13.2) | 414 (11.1) |
| 2 | 7113 (46.8) | 1809 (47.2) | 1800 (46.7) | 1774 (47.0) | 1730 (46.4) |
| ≥3 | 3443 (22.7) | 885 (23.1) | 892 (23.1) | 820 (21.7) | 846 (22.7) |
| Age at first birth |  |  |  |  |  |
| Nulliparous | 2782 (18.3) | 694 (18.1) | 671 (17.4) | 681 (18.0) | 736 (19.8) |
| <25 | 4694 (30.9) | 1422 (37.1) | 1265 (32.8) | 1101 (29.2) | 906 (24.3) |
| 25–29 | 4870 (32.1) | 1147 (29.9) | 1244 (32.3) | 1239 (32.8) | 1240 (33.3) |
| ≥30 | 2835 (18.7) | 567 (14.8) | 675 (17.5) | 752 (19.9) | 841 (22.6) |
| Age at menopause |  |  |  |  |  |
| <45 | 1792 (12.4) | 537 (14.7) | 462 (12.7) | 399 (11.1) | 394 (11.1) |
| 45–49 | 3296 (22.8) | 792 (21.7) | 833 (22.8) | 866 (24.0) | 805 (22.7) |
| 50–54 | 6856 (47.5) | 1697 (46.6) | 1690 (46.3) | 1699 (47.1) | 1770 (50.0) |
| ≥55 | 2383 (16.5) | 583 (16.0) | 630 (17.3) | 620 (17.2) | 550 (15.5) |
| Unknown | 109 (0.8) | 34 (0.9) | 32 (0.9) | 23 (0.6) | 20 (0.6) |

Data are presented as median (interquartile range) for continuous variables and count (percent) for categorical variables.
Unknown age at menopause represents structurally unavailable age at menopause information; ordinary missing values were excluded from the descriptive distribution.
BMI, body mass index; CRF, cardiorespiratory fitness; PRS, polygenic risk score; $\dot{V}O_{2\max}$ , maximal oxygen uptake

### Association between PRS and Incident Breast Cancer

The association between PRS and incident breast cancer is shown in Supplemental Table 3 (Supplemental Digital Content 1). After adjustment for covariates (Model 2), higher PRS was associated with progressively higher risk of incident breast cancer (HR per 1-SD increase 1.78, 95% CI 1.63 to 1.93; *P* < 0.001). Restricted cubic spline analyses further demonstrated an approximately linear dose–response relationship between PRS and breast cancer risk (*P* for nonlinearity = 0.274, Supplemental Figure 1, Supplemental Digital Content 1). Nelson–Aalen curves were consistent with the Cox regression results, showing progressively greater cumulative hazard across higher PRS quartiles (Figure 2A). Age-stratified analysis showed that the association between PRS and incident breast cancer was stronger in women aged ≥60 years than in those aged 40–59 years (Supplemental Table 3 and Supplemental Figure 2, Supplemental Digital Content 1). Similar associations were observed for breast cancer-specific mortality (Supplemental Table 4, Supplemental Digital Content 1).

**Figure 2.**
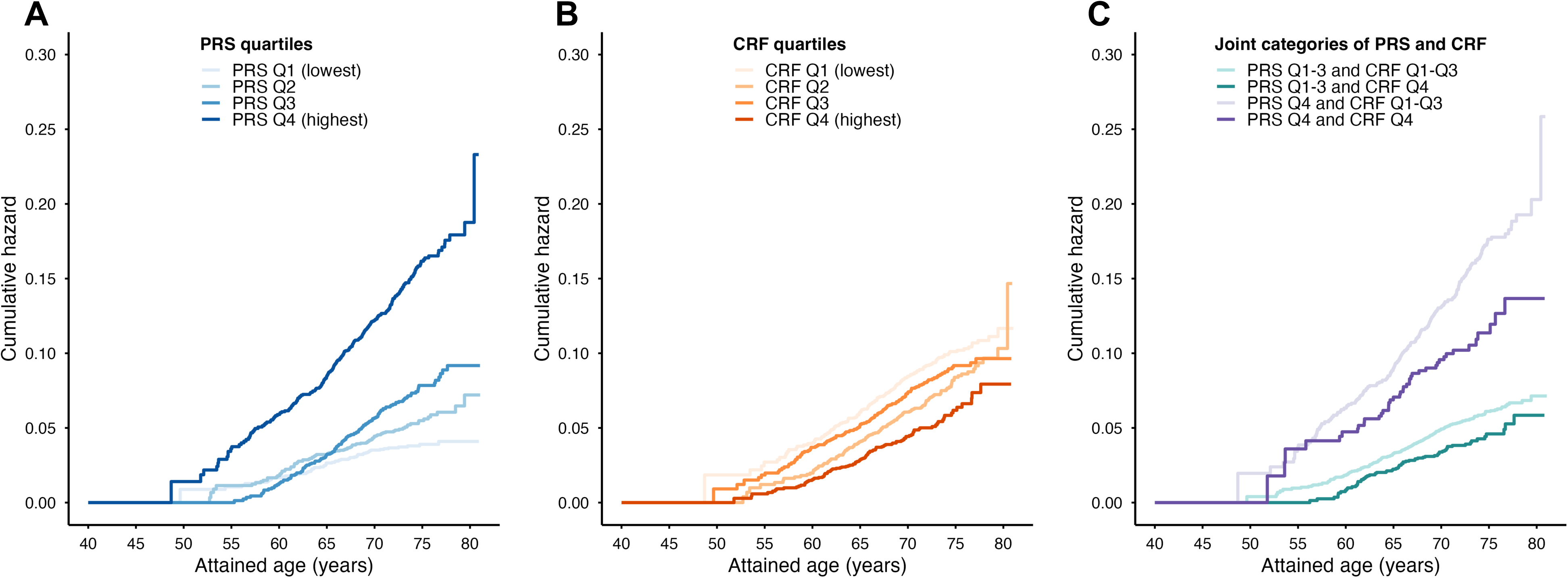
Nelson–Aalen cumulative hazard curves for incident breast cancer according to (A) PRS quartiles, (B) CRF quartiles, and (C) joint categories of PRS and CRF.

### Association between CRF and Incident Breast Cancer

The association between CRF quartiles and incident breast cancer is shown in Table 2. In Model 2, CRF was inversely associated with incident breast cancer (HR per 1-MET increase 0.91, 95% CI 0.86 to 0.97, *P* = 0.004). Further adjustment for body fat percentage in Model 3 did not substantially change the results (HR 0.92, 95% CI 0.85 to 0.99, *P* = 0.028). Restricted cubic spline analyses were consistent with these findings and supported an approximately linear inverse dose–response relationship between CRF and breast cancer risk (*P* for nonlinearity = 0.298 in Model 2, Supplemental Figure 3, Supplemental Digital Content 1). Cumulative hazard curves were consistent with the Cox regression results, with the lowest cumulative hazard observed in the highest CRF quartile (Figure 2B). Similar trends were observed in age- and BMI-stratified analyses (Supplemental Table 5, Supplemental Figures 4 and 5, Supplemental Digital Content 1). However, a significant linear association was observed only in women aged ≥60 years (HR 0.89, 95% CI 0.82 to 0.96, *P* = 0.004) and those with BMI ≥25 kg/m^2^ (HR 0.89, 95% CI 0.81 to 0.97, *P* = 0.012), but not in those aged 40–59 years (HR 0.95, 95% CI 0.86 to 1.06, *P* = 0.352) or those with BMI <25 kg/m^2^ (HR 0.97, 95% CI 0.88 to 1.08, *P* = 0.611). In the sensitivity analysis excluding breast cancer cases diagnosed within the first 2 years of follow-up, the inverse association was attenuated and was no longer statistically significant, although the direction of association remained consistent (Supplemental Tables 6 and 7, Supplemental Digital Content 1). Results were also consistent in the competing-risk analysis using Fine–Gray models, with higher CRF associated with a lower subdistribution hazard of incident breast cancer (Supplemental Tables 8 and 9, Supplemental Digital Content 1). The association between CRF and breast cancer-specific mortality was not statistically significant (Supplemental Table 10, Supplemental Digital Content 1).

**Table 2.** Association between CRF and incident breast cancer.

| CRF | *n* | Events | Incidence rate<br>(per 1000 PY) | Model 1 HR<br>(95% CI) | Model 2 HR<br>(95% CI) | Model 3 HR<br>(95% CI) |
| --- | --- | --- | --- | --- | --- | --- |
| Q1 | 3835 | 148 | 3.7 | 1.00 (ref) | 1.00 (ref) | 1.00 (ref) |
| Q2 | 3859 | 147 | 3.7 | 0.98 (0.78, 1.24) | 0.97 (0.77, 1.22) | 0.99 (0.78, 1.25) |
| Q3 | 3776 | 144 | 3.7 | 0.98 (0.78, 1.24) | 0.95 (0.75, 1.20) | 0.98 (0.76, 1.26) |
| Q4 | 3729 | 108 | 2.8 | 0.75 (0.58, 0.96) | 0.72 (0.56, 0.93) | 0.76 (0.56, 1.02) |
| Per 1-MET increase |  |  |  | 0.92 (0.87, 0.98)<br>*P* = 0.009 | 0.91 (0.86, 0.97)<br>*P* = 0.004 | 0.92 (0.85, 0.99)<br>*P* = 0.028 |

Model 1 used attained age as the underlying time scale. Model 2 was adjusted for family history of breast cancer, Townsend deprivation index, alcohol consumption frequency, smoking status, hormone replacement therapy use, age at menarche, number of live births, age at first birth among parous women, and age at menopause. Model 3 was additionally adjusted for body fat percentage.
CI, confidence interval; CRF, cardiorespiratory fitness; HR, hazard ratio; MET, metabolic equivalent of task; PY, person-years; Ref, reference

### Joint Associations of CRF and PRS with Incident Breast Cancer

Joint associations of CRF and PRS quartiles with incident breast cancer are shown in Figure 3. Women with high PRS (Q4) and low-to-moderate CRF (Q1–Q3) exhibited the highest risk of breast cancer in the overall sample (HR 2.59, 95% CI 2.14 to 3.12, *P* < 0.001 in Model 2). Although no significant multiplicative interaction between CRF and PRS was detected (interaction HR 0.75, 95% CI 0.49 to 1.16, *P* for interaction = 0.201), additive interaction analyses demonstrated a significant negative RERI in the overall sample (RERI −0.79, 95% CI −1.47 to −0.10, *P* = 0.025). In age-stratified analyses, this additive interaction was particularly pronounced among women aged ≥60 years (RERI −1.14, 95% CI −1.95 to −0.33, *P* = 0.006), whereas no significant interaction was observed among women aged 40–59 years (RERI 0.04, 95% CI −1.26 to 1.33, *P* = 0.957). In this older age group, women with high PRS and high CRF did not show a statistically significant increase in breast cancer risk compared with the reference group (HR 1.30, 95% CI 0.83 to 2.03, *P* = 0.255). The RERI estimates were similar across BMI strata (BMI <25 kg/m²: RERI −1.10, 95% CI −2.44 to 0.24, *P* = 0.106; BMI ≥25 kg/m²: RERI −1.10, 95% CI −2.06 to −0.14, *P* = 0.025), although the 95% CI included the null among women with BMI <25 kg/m². Among women with overweight or obesity, those with high PRS and high CRF did not show a statistically significant increase in breast cancer risk compared with the reference group (HR 1.00, 95% CI 0.47 to 2.13, *P* = 0.995). Further adjustment for body fat percentage in Model 3 did not substantially change these results (Supplemental Table 11, Supplemental Digital Content 1). In the sensitivity analysis excluding breast cancer cases diagnosed within the first 2 years of follow-up, the overall pattern of joint associations was similar, although the additive interaction was attenuated and no longer statistically significant in the overall and age ≥60-year analyses (Supplemental Table 12, Supplemental Digital Content 1). Competing-risk analyses using Fine–Gray models also yielded results broadly consistent with the primary Cox analyses for the joint associations and interactions between CRF and PRS (Supplemental Table 13, Supplemental Digital Content 1). Cumulative hazard curves according to joint CRF and PRS categories further supported these findings. Among women with high PRS, the cumulative hazard curves were largely overlapping between CRF groups before age 65 years but began to diverge thereafter, with higher CRF associated with a slower accumulation of breast cancer risk (Figure 2C).

**Figure 3.**
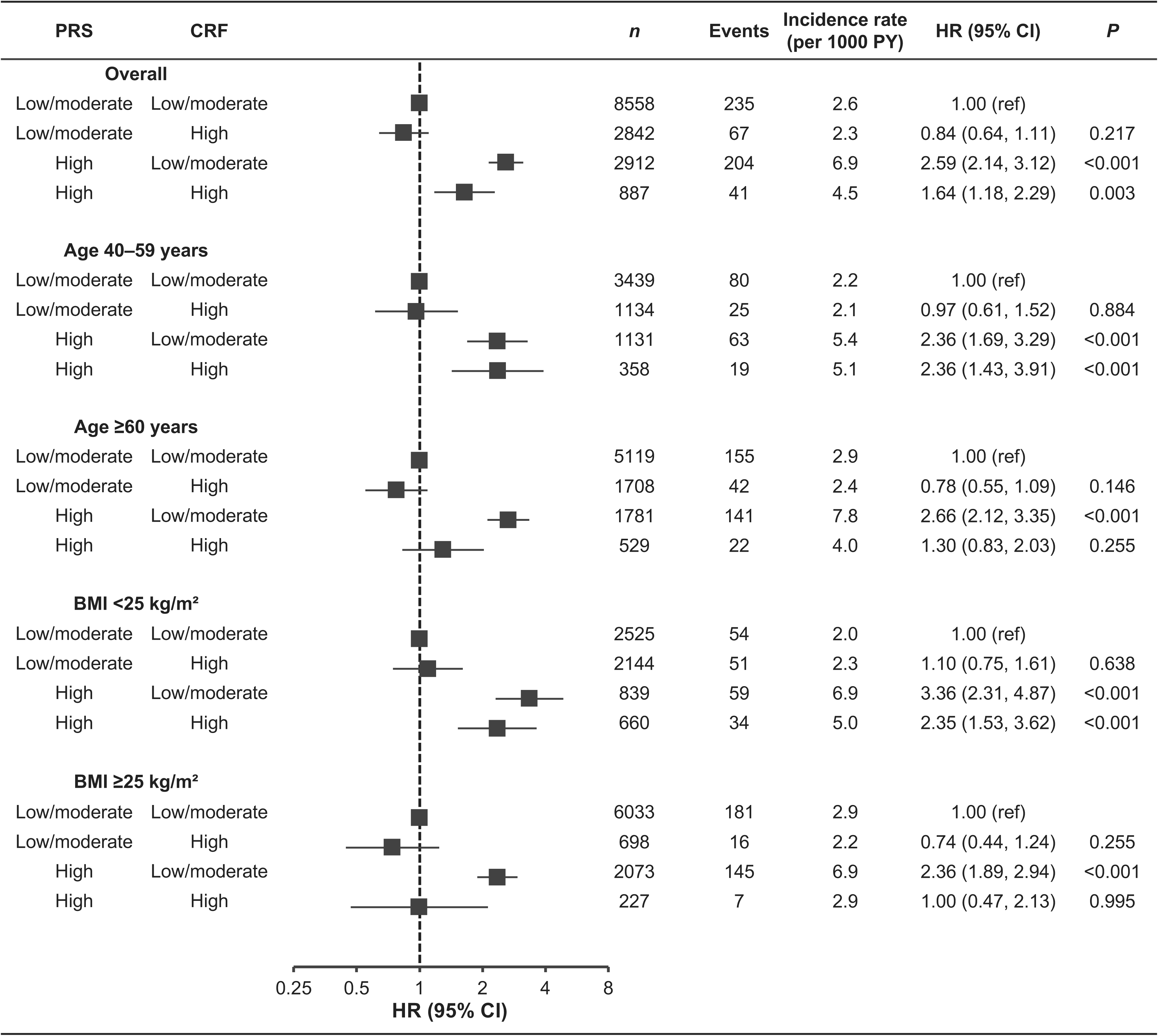
Joint associations and interaction of CRF and PRS with incident breast cancer. CRF and PRS were dichotomized into low/moderate (Q1–Q3) and high (Q4) categories. Models were adjusted for family history of breast cancer, Townsend deprivation index, alcohol consumption frequency, smoking status, hormone replacement therapy use, age at menarche, number of live births, age at first birth, and age at menopause.

## Discussion

In this prospective cohort study of postmenopausal women from the UK Biobank, CRF was inversely associated with incident breast cancer, whereas breast cancer PRS was positively associated with breast cancer risk. Higher CRF was associated with attenuation of excess breast cancer risk conferred by high polygenic risk on the additive scale, and this attenuation was more pronounced among those aged ≥60 years. In contrast, attenuation of excess breast cancer risk was not evident among women aged 40–59 years. The RERI estimates were similar across BMI strata, although the 95% CI included the null among women with BMI <25 kg/m². No clear association was observed for breast cancer–specific mortality. These findings suggest that higher CRF was associated with attenuation of excess breast cancer risk conferred by elevated polygenic risk in postmenopausal women, particularly among older women.

### Comparison with Previous Studies

In the present study, higher CRF was associated with a lower risk of breast cancer overall, consistent with previous prospective evidence (12). In BMI-stratified analyses, the inverse association between CRF and breast cancer risk was observed among women with overweight or obesity but not in those with BMI <25 kg/m². A prospective analysis of UK Biobank similarly reported that higher CRF was associated with a lower breast cancer risk primarily among women with overweight or obesity (11). We also observed that the inverse association between CRF and breast cancer risk was most evident among women aged ≥60 years. These findings suggest that CRF may play a particularly important role among older women and those with overweight or obesity. Notably, the inverse association between CRF and breast cancer risk remained after additional adjustment for body fat percentage, suggesting that the observed association was not explained solely by adiposity.

Our findings are also consistent with previous studies reporting that favorable lifestyle behaviors, including higher levels of physical activity, may attenuate the excess breast cancer risk associated with elevated polygenic risk (17). In the present study, multiplicative interaction between CRF and PRS was not statistically significant. Nevertheless, additive interaction analyses together with the joint association analyses suggested that higher CRF was associated with attenuation of excess breast cancer risk conferred by high PRS. From a public health perspective, additive interaction may be particularly informative because it reflects the absolute excess risk attributable to the combined effects of two risk factors (18) and therefore has greater relevance for identifying populations most likely to benefit from preventive strategies. To our knowledge, no previous prospective studies have investigated the joint associations and potential interaction between objectively assessed CRF and genetic susceptibility in relation to incident breast cancer. The present findings extend existing evidence by indicating that objectively assessed CRF may also be associated with attenuation of the excess breast cancer risk conferred by elevated polygenic risk.

### Biological Interpretation

The finding that higher CRF was associated with attenuation of excess breast cancer risk conferred by high polygenic risk among women aged ≥60 years is biologically plausible. Aging is accompanied by immune senescence (19), hormonal changes including increased peripheral production of estrogen in adipose tissue (20), and accumulated cellular damage (21), which may amplify the phenotypic manifestation of genetic risk. Together, these processes may create a protumorigenic environment. Higher CRF is associated with improved metabolic regulation (22), reduced systemic inflammation (23), and enhanced immune surveillance (24), which may help limit tumor development. Among women aged ≥60 years, where genetic susceptibility may be more strongly expressed with advancing age, physiological resilience indexed by CRF may therefore exert a more apparent influence.

The absence of clear attenuation among women aged 40–59 years warrants cautious interpretation. Breast cancer etiology in relatively younger postmenopausal women may be more strongly shaped by life-course reproductive factors (25) and cumulative hormonal exposures established earlier in life (26), potentially diminishing the relative contribution of CRF. In addition, the relatively small number of breast cancer cases in this subgroup may have limited statistical power to detect modest associations. Further large-scale prospective studies are needed to confirm these subgroup-specific findings.

### Strengths and Limitations

This study has several strengths. The prospective design enabled robust assessment of incident breast cancer. The use of objectively estimated CRF together with a breast cancer PRS enabled evaluation of the joint associations of fitness and genetic susceptibility. The assessment of additive interaction offers clinically and public health–relevant insight into risk modification.

Several limitations should be considered when interpreting these findings. First, CRF was estimated rather than directly measured using gas exchange analysis, which may have introduced non-differential measurement error and attenuated associations. However, the UK Biobank submaximal cycle test has been validated for estimating CRF in population studies (15). Second, CRF was assessed only at baseline, and changes over time were not captured, potentially limiting assessment of long-term exposure effects. Third, the analytic sample was restricted to women of European genetic ancestry, all of whom were classified as White British based on self-reported ethnic background, which may limit generalizability to other populations. Fourth, residual confounding may remain despite adjustment for established breast cancer risk factors. Fifth, the number of incident breast cancer cases was relatively small in some subgroup and joint CRF–PRS categories. In particular, the small number of events in the high-CRF categories among women with BMI ≥25 kg/m² resulted in wide CIs and limited precision of the joint association and RERI estimates. Accordingly, the BMI-stratified joint association and interaction findings should be considered exploratory and hypothesis-generating and interpreted cautiously. Sixth, only participants completing the submaximal fitness test were included, which may introduce selection bias and limit generalizability. Finally, as an observational study, causal inference cannot be established, and the findings require replication in independent cohorts.

### Conclusions

In this prospective cohort of postmenopausal women, higher CRF was associated with a lower breast cancer risk and attenuation of excess breast cancer risk conferred by elevated polygenic risk, particularly among postmenopausal women aged ≥60 years. These findings suggest that maintaining higher CRF may be particularly beneficial among postmenopausal women with elevated polygenic risk and support the potential role of CRF in risk-stratified breast cancer prevention strategies.

## Supporting information

Supplemental Digital Content

## Data Availability

The data used in this study are available from the UK Biobank under application number 185057. Researchers can apply for access to the UK Biobank resource through the UK Biobank Access Management System (https://www.ukbiobank.ac.uk/).

https://www.ukbiobank.ac.uk/enable-your-research/apply-for-access

## Acknowledgements

This research was conducted using the UK Biobank Resource under application number 185057. The authors thank the UK Biobank participants and staff for their valuable contributions. This study was supported by the National Natural Science Foundation of China (No. 32371228), the Key Research and Development Program of Shaanxi Province (No. 2025SF-YBXM-367), and the Fundamental Research Funds of Xi’an Jiaotong University (xtr052023011).

The results of the present study are presented clearly, honestly, and without fabrication, falsification, or inappropriate data manipulation. The results of the present study do not constitute endorsement by the ACSM.

## Supplemental Digital Contents

Supplemental Digital Contents 1: Supplemental_Digital_Content.doc

## References

1. Bray F, Laversanne M, Sung H, et al. Global cancer statistics 2022: GLOBOCAN estimates of incidence and mortality worldwide for 36 cancers in 185 countries. CA Cancer J Clin. 2024;74(3):229–63.

2. £ukasiewicz S, Czeczelewski M, Forma A, Baj J, Sitarz R, Stanisławek A. Breast cancer—epidemiology, risk factors, classification, prognostic markers, and current treatment strategies—An updated review. Cancers (Basel). 2021;13(17).

3. Turnbull C, Rahman N. Genetic predisposition to breast cancer: Past, present, and future. Annu Rev Genomics Hum Genet. 2008;9:321–45.

4. Michailidou K, Lindström S, Dennis J, et al. Association analysis identifies 65 new breast cancer risk loci. Nature. 2017;551(7678):92–4.

5. Michailidou K, Beesley J, Lindstrom S, et al. Genome-wide association analysis of more than 120,000 individuals identifies 15 new susceptibility loci for breast cancer. Nat Genet. 2015;47(4):373–80.

6. Mavaddat N, Michailidou K, Dennis J, et al. Polygenic Risk Scores for Prediction of Breast Cancer and Breast Cancer Subtypes. Am J Hum Genet. 2019;104(1):21–34.

7. Yanes T, Young MA, Meiser B, James PA. Clinical applications of polygenic breast cancer risk: A critical review and perspectives of an emerging field. Breast Cancer Research [Internet]. 2020;22(1) doi:10.1186/s13058-020-01260-3.

8. Ross R, Blair SN, Arena R, et al. Importance of Assessing Cardiorespiratory Fitness in Clinical Practice: A Case for Fitness as a Clinical Vital Sign: A Scientific Statement from the American Heart Association. Circulation. 2016;134(24):e653–99.

9. Lang JJ, Prince SA, Merucci K, et al. Cardiorespiratory fitness is a strong and consistent predictor of morbidity and mortality among adults: an overview of meta-analyses representing over 20.9 million observations from 199 unique cohort studies. Br J Sports Med. 2024;58(10):556–66.

10. Kodama S, Saito K, Tanaka S, et al. Cardiorespiratory Fitness as a Quantitative Predictor of All-Cause Mortality and Cardiovascular Events in Healthy Men and Women A Meta-analysis. JAMA. 2009;301(19):2024–35.

11. Christensen RAG, Knight JA, Sutradhar R, Brooks JD. Association between estimated cardiorespiratory fitness and breast cancer: A prospective cohort study. Br J Sports Med. 2023;57(19):1238–47.

12. Katsaroli I, Sidossis L, Katsagoni C, et al. The Association between Cardiorespiratory Fitness and the Risk of Breast Cancer in Women. Med Sci Sports Exerc. 2024;56(6):1134–9.

13. Sudlow C, Gallacher J, Allen N, et al. UK Biobank: An Open Access Resource for Identifying the Causes of a Wide Range of Complex Diseases of Middle and Old Age. PLoS Med [Internet]. 2015;12(3) doi:10.1371/journal.pmed.1001779.

14. UK Biobank. UK Biobank Cardio Assessment Manual Version 1.0. https://biobank.ctsu.ox.ac.uk/crystal/crystal/docs/Cardio.pdf. [date unknown];

15. Gonzales TI, Westgate K, Strain T, et al. Cardiorespiratory fitness assessment using risk-stratified exercise testing and dose–response relationships with disease outcomes. Sci Rep [Internet]. 2021;11(1) doi:10.1038/s41598-021-94768-3.

16. Thompson DJ, Wells D, Selzam S, et al. A systematic evaluation of the performance and properties of the UK Biobank Polygenic Risk Score (PRS) Release. PLoS One [Internet]. 2024;19(9 September) doi:10.1371/journal.pone.0307270.

17. Arthur RS, Wang T, Xue X, Kamensky V, Rohan TE. Genetic factors, adherence to healthy lifestyle behavior, and risk of invasive breast cancer among women in the UK biobank. J Natl Cancer Inst. 2020;112(9):893–901.

18. Knol MJ, VanderWeele TJ. Recommendations for presenting analyses of effect modification and interaction. Int J Epidemiol. 2012;41(2):514–20.

19. Wang L, Tang D. Immunosenescence promotes cancer development: from mechanisms to treatment strategies. Cell Communication and Signaling [Internet]. 2025;23(1) doi:10.1186/s12964-025-02082-6.

20. Bhardwaj P, Au CMC, Benito-Martin A, et al. Estrogens and breast cancer: Mechanisms involved in obesity-related development, growth and progression. Journal of Steroid Biochemistry and Molecular Biology. 2019;189:161–70.

21. Gladyshev VN, Kritchevsky SB, Clarke SG, et al. Molecular damage in aging. Nat Aging. 2021;1(12):1096–106.

22. Myers J, Kokkinos P, Nyelin E. Physical activity, cardiorespiratory fitness, and the metabolic syndrome. Nutrients [Internet]. 2019;11(7) doi:10.3390/nu11071652.

23. Kullo IJ, Khaleghi M, Hensrud DD. Markers of inflammation are inversely associated with VO_2_ max in asymptomatic men. J Appl Physiol. 2007;102:1374–9.

24. Lyu DW. Immunomodulatory effects of exercise in cancer prevention and adjuvant therapy: a narrative review. Front Physiol. 2023;14:1292580.

25. Colditz GA, Bohlke K, Berkey CS. Breast cancer risk accumulation starts early: Prevention must also. Breast Cancer Res Treat. 2014;145(3):567–79.

26. Clavel-Chapelon F, Gerber M. Reproductive factors and breast cancer risk. Do they differ according to age at diagnosis? Breast Cancer Res Treat. 2002;72:107–15.

